# Multispecialty multidisciplinary input into comorbidities in heart failure reduces hospitalisation and clinic attendance

**DOI:** 10.1101/2022.01.31.22270113

**Authors:** Hani Essa, Lauren Walker, Kevin Mohee, Emeka Oguguo, Homeyra Douglas, Matthew Kahn, Archana Rao, Julie Bellieu, Justine Hadcroft, Nick Hartshorne-Evans, Janet Bliss, Asan Akpan, Christopher Wong, Daniel J Cuthbertson, Rajiv Sankaranarayanan

## Abstract

**Aims:** Heart failure (HF) is associated with multiple co-morbidities which independently influence response to treatment as well as outcomes. This retrospective observational study (January 2020-June 2021) analysed the impact of monthly virtual HF multi-specialty multi-disciplinary team (MDT) meetings to address the management of associated comorbidities and thereby upon provision, cost of care and HF outcomes.

**Methods:** Patients acted as their own controls, with outcomes compared for equal periods (for each patient) pre versus post-MDT meeting. The MDT comprised of HF cardiologists (primary, secondary, tertiary care), HF specialist nurses (hospital, community), nephrologist, endocrinologist, palliative care specialist, chest physician, pharmacist, clinical pharmacologist and geriatrician. Outcome measures were 1) all-cause hospitalisations, 2) outpatient clinic attendances, and 3) cost.

**Results:** 334 patients (mean age 72.5±11 years) were discussed virtually through MDT meetings and follow-up duration was 13.9 ± 4 months. The mean age-adjusted Charlson Co-morbidity Index was 7.6 ± 2.1 and Rockwood Frailty Score was 5.5 ± 1.6. The mean number of clinic attendances prevented was 1.6 ± 0.4. The total cost of funding monthly meetings for the duration of the study was £32400 and the 64 clinic appointments generated cost £9600. The MDT meetings prevented 534 clinic appointments (cost saving £80,100) and reduced all-cause hospitalisations (pre-MDT meeting 1.1±0.4 vs. 0.6±0.1 post-MDT meeting; p<0.001), reduction of 1586 hospital bed-days and cost-savings of £634,400. The total cost-saving to the healthcare system was £672,500.

**Conclusion:** The HF multispecialty virtual MDT model provides integration of care across all tiers of healthcare for HF management and a holistic approach addressing associated co-morbidities. This approach can reduce the need for out-patient attendances and all-cause hospitalisations, leading to significant cost-savings.

**Key questions:** *What is already known about this subject?:* Heart failure is associated with several co-morbid health conditions (multi-morbidity) which independently influence outcomes as well as response to treatment.

*What does this study add?:* This study assesses the impact of multispecialty multi-morbidity input into the management of co-morbidities and thereby the effect upon all-cause outcomes.

*How might this impact on clinical practice?:* Results of this study illustrate that multi-speciality management of comorbidities associated with heart failure, may not only improve all-cause outcomes but could also prove to be cost-beneficial.

## Introduction

Heart failure (HF) is a complex clinical syndrome, representing the final common pathway of many different pathological processes and associated with high mortality and frequent hospital admissions^1^. There are an estimated 60 million cases of HF worldwide^2^. This global burden is expected to increase with an aging population, with 80% of hospitalisations occurring in those aged >65 years^3^. Significantly, hospitalisation confers a poor long term prognosis with a 5-year mortality of 75%^4^.

HF is frequently characterised by multi-morbidity with patients suffering from many other comorbid conditions (long-term conditions) that can influence the management and adversely affect outcomes. These include other cardiovascular diseases such as ischaemic heart disease (IHD), atrial fibrillation (AF) and hypertension and non-cardiovascular comorbidities such as chronic kidney disease (CKD) (41%), anaemia (37%), diabetes mellitus (type 1 or type 2) (DM-21%), chronic obstructive pulmonary disease (COPD) (24%), a burden of polypharmacy and frailty ^5, 6^. Most HF patients have at least one comorbidity^6, 7^ with up to 40% of patients with HF with ≥5 co-morbidities (this group contributed to >80% of the overall in-hospital stay)^8^. The number of comorbidities is higher amongst patients with heart failure with preserved ejection fraction (HFpEF) than in patients with heart failure with reduced ejection fraction (HFrEF) and with a more profound impact upon outcomes ^9-13^. In addition, non-cardiac comorbidities independently affect response to treatment, influence disease severity and HF outcomes such as hospitalisation, quality of life and mortality ^7, 12-19^. The burden of comorbidities is also progressively increasing with time^13, 20^.

This level of multi-morbidity and complexity can be most effectively managed by a multidisciplinary team (MDT), an approach endorsed by both the European Society of Cardiology (ESC) (Class 1A recommendation)^21^ and the American Heart Association (AHA) (Class 1B recommendation)^21^. However, there is little detail in the guidelines about the composition or remit of such an MDT. There is also a dearth of evidence currently regarding whether management of comorbidities by relevant specialists in consultation with HF specialists can improve HF outcomes.

The aim of this study is to assess the impact of multi-specialty input on the optimisation of comorbidities, all-cause hospitalisations, out-patient clinic attendances and mortality in patients with heart failure.

## Methods

### Study settings and time period

This study complies with the Declaration of Helsinki and was approved as a service evaluation project by the Research Governance Committee (Institutional Review Board) at Liverpool University Hospitals NHS Foundation Trust and Liverpool Health Partners. It was conducted as an observational cohort study in a British University teaching hospital, looking at the impact of a multispecialty HF MDT meeting on care models and outcomes in a HF population. We studied outcomes of our patients with HF at monthly virtual multispecialty meetings from January 2020 to June 2021 and followed up for equal amounts of time pre and post recruitment. We followed the STROBE guidelines (The Strengthening the Reporting of Observational Studies in Epidemiology) for reporting observational studies^22^.

### MDT meeting format

Since January 2020, the Liverpool Multi-specialty regional HF MDT meetings have been funded by Liverpool Single Services Cardio-respiratory Group (from Liverpool Clinical Commissioning Group) and Liverpool University Hospitals NHS Foundation Trust. The multi-specialty meeting is conducted monthly via video-conferencing. Sources of referral include community heart failure teams, hospital HF teams (secondary and tertiary care) and other specialty teams, using a dedicated referral form (Supplementary material). The multispecialty multidisciplinary team consist of HF cardiologists from the community, secondary care and tertiary care, heart failure specialist nurses from the community and hospital, nephrologist, endocrinologist, palliative care specialist, chest physician, geriatrician, pharmacist and pharmacologist. Individualised discussions at the multispecialty MDT meeting include optimisation of HF therapies, assessment of frailty, comorbidities, cardio-renal metabolic status, specialist input into any complex chronic respiratory pathologies, need for falls risk assessment, cognitive dysfunction, rationalisation of polypharmacy burden (particularly anti-cholinergic burden), medication compliance and need for advanced care planning discussions or community palliative care where appropriate. Recommendations are made as consensus from the multispecialty meeting. These are then conveyed electronically or through post to the referrers, the electronic record of each discussed patient is also updated with these recommendations and the MDT consensus discussed with each patient.

### Demographic and clinical details

We collected demographic data, medication history, co-morbidities such as diabetes mellitus, hypertension, ischaemic heart disease (IHD), chronic obstructive pulmonary disease (COPD) or a pre-existing malignant condition. Furthermore we computed the Charlson Co-morbidity Index (CCI)^23^ and the Rockwood clinical frailty score^24^. Patients were followed up for equal amounts of time pre and post MDT meeting, so “each patient acted as their own control”.

### Outcome measures

Outcome measures assessed were all-cause hospitalisations and outpatient clinic attendances for an equal period of time pre and post MDT meeting. We also assessed other measures such as advanced HF management (device therapy, transplant referrals), referrals to integrated palliative care services, burden of polypharmacy and all-cause mortality.

### Statistical analysis

For the descriptive statistics of our patient population, we represented continuous variables as means with standard deviations or medians with interquartile ranges in the case of non-parametric data. Categorical variables were represented with percentages and analysed using Chi-squared test. Statistical comparisons were made for parametric data using Student’s T test and for nonparametric data using Mann-Whitney test. Statistical analysis was carried out using the Statistical Package for the Social Sciences (SPSS 23.0, IBM Corp., Armonk, New York, USA).

## Results

### Demographic details

334 patients were discussed through virtual MDT meetings from January 2020 to June 2021. We collated baseline patient characteristics as illustrated in Table 1. This service model continued uninterrupted during the COVID-19 pandemic. The mean age of patients discussed was 72.5±11 years and follow-up duration was 13.9 ± 4 months. 43% of the patients discussed were female and 45% of patients had a diagnosis of HFpEF.

**Table 1.** Patient Characteristics

| **Patient Characteristics** | **N =334** |
| --- | --- |
| Age | 72.5±11 years |
| Male : Female | 57%:43% |
| HFrEF / HFmrEF | 55% |
| HFpEF | 45% |
| Ischaemic Heart Disease | 37% |
| Hypertension | 49% |
| Diabetes Mellitus | 41% |
| Atrial Fibrillation | 38% |
| Valvular Heart Disease | 18% |
| COPD / Interstitial Lung Disease | 31% |
| CKD | 53% |
| Cerebrovascular disease (CVA or previous TIA) | 13% |
| Cancer | 7% |
| Dementia | 8% |
| Charlson Comorbidity Index | 7.6 ± 2.1 |
| Rockwood Frailty Score | 5.5 ± 1.6 |

The mean age-adjusted Charlson Co-morbidity Index was 7.6 ± 2.1 and Rockwood Frailty Score was 5.5 ± 1.6. There was significant polypharmacy burden as indicated by the mean number of medications of 12.1± 3.9 amongst the whole cohort.

### Clinical recommendations

Consensus recommendation was made from the MDT regarding medication optimisation such as addition of sodium glucose-like co-transporter-1 (SGLT2inhibitor) in patients with HF, diabetes or CKD. Appropriate de-prescribing of polypharmacy was also performed to reduce the anticholinergic burden (mean number of medications reduced -1.4 ± 1) and 9% of patients were referred to the falls assessment clinic. (53/334 (16%) patients underwent internal cardiovertor-defibrillator (ICD) or cardiac resynchronisation therapy (CRT) pacemaker implant as a result of consensus recommendations from the multispecialty MDT and 11/334 (3%) patients were referred for transplant assessment. (16/334) 5% of patients were referred to dialysis assessment clinics and 10 patients were instituted on dialysis.

37/334 (11%) patients required community palliative care, 9 of these patients required admission to a hospice and 11 patients required deactivation of their ICD as a part of advanced care planning palliative care. Overall mortality of this cohort during follow-up was 21% (70/334).

### Hospitalisations

As shown in figure 1, the number of all-cause hospitalisations was reduced significantly (pre-MDT 1.1±0.4 vs.0.6±0.1 post MDT; p<0.001), leading to a saving of 1586 bed-days and estimated cost-saving £634,400 (average cost per bed day £400^25^).

**Figure 1.**
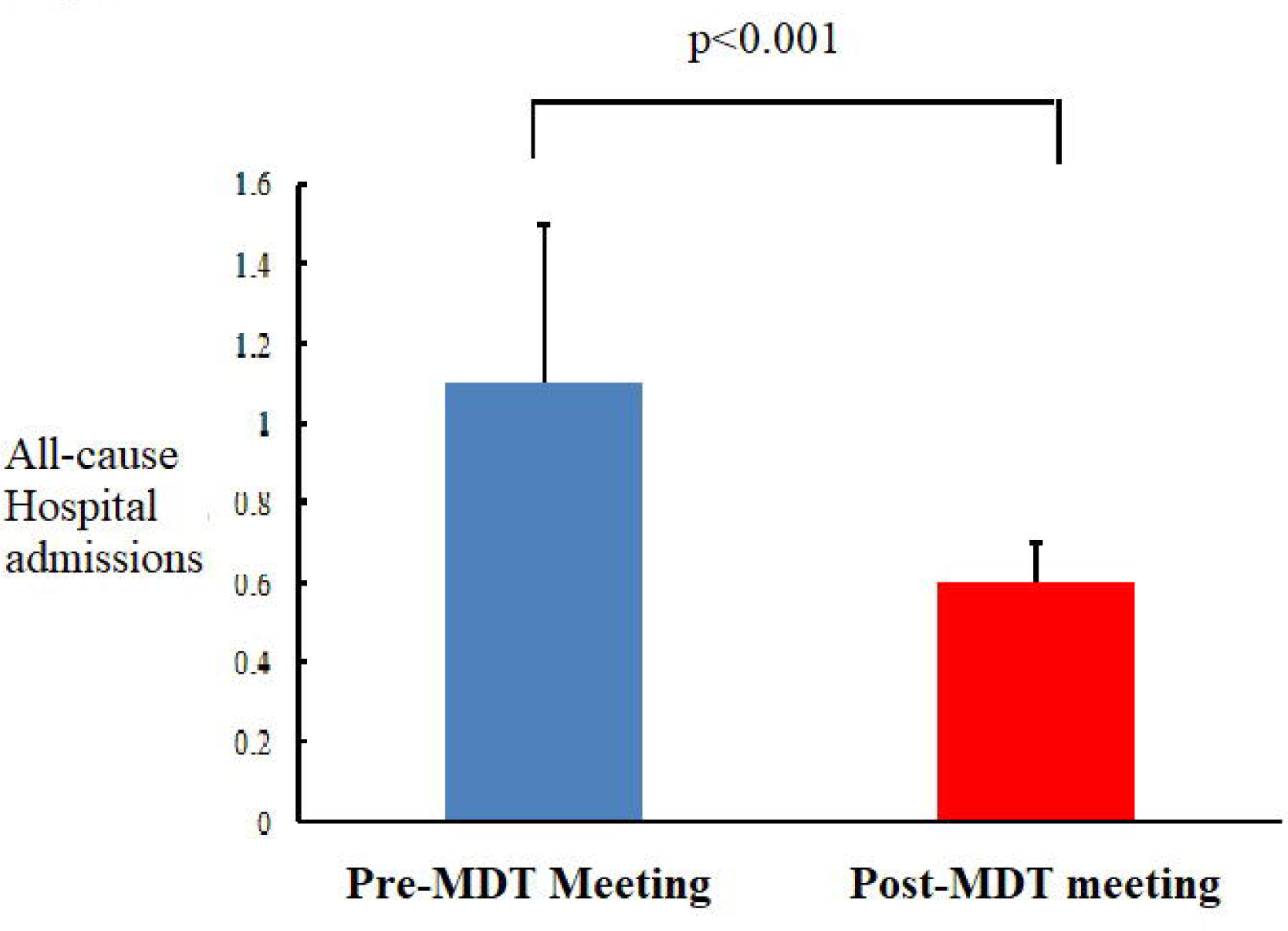
Comparison of all-cause hospitalisations pre and post-multispeciality MDT meeting

### Out-patient attendances

The number of outpatient clinic attendances prevented 534. This reduced inconvenience to patients and also potentially saved patients money. This includes avoidance of need for travel as well as waiting time in clinic (average 51.3 minutes^26^, transport and parking costs (average saving of £5.52 per patient per appointment by car and £4.60 by bus)^27^. There was also the added positive environmental impact through carbon footprint reduction (in this study 554 kgCO2).

### Economic analysis

We also performed an economic analysis of the impact of the virtual multispecialty MDT. This is also illustrated in Table 2. The total cost of funding the multispecialty MDT meeting was 0.5 programmed activity (PA) NHS rates^28^ per speciality (£1800 per meeting and £32400 for the duration of the study (Jan 2020 – Jun 2021). 64 clinic appointments they generated cost an estimated £9600 (£150 per outpatient clinic^25^). However, the MDT meetings also prevented 534 clinic appointments (cost saving £80,100). As shown in Table 2, the total saving to the healthcare system was £672,500.

**Table 2.** Economic analysis- healthcare savings

|  | **Expenditure** | **Saving** |
| --- | --- | --- |
| **Funding of specialties for MDT input** | £32400 |  |
| **Reduction in hospitalisations post MDT** |  | 1.1 pre MDT to 0.6 post (3490-1904= saving of 1586 bed-days =£634,400) |
| **Outpatient clinic visits post MDT** | 64 generated<br>= £9600 | 534 clinic appointments prevented (cost saving £80,100) |
|  | Total<br>expenditure<br>£42000 | Total saving<br>£714,500 |
| **Total saving to the healthcare system =**<br>£672,500 |  |  |

## Discussion

Several previous studies have shown the co-existence and adverse impact of multi-morbidity in patients with HF. However, this is the first study to demonstrate that multispecialty, multidisciplinary management of comorbidities with integration of community, secondary and tertiary care HF specialists, is associated with reduced all-cause hospitalisations and outpatient clinic attendances. The MDT also optimised the care of multiple comorbidities and facilitated timely involvement of relevant specialties.

It is important to focus on the management of multi-morbidity along with the management of HF itself. Other studies have demonstrated that HF contributes to a smaller proportion of the burden of hospitalisation (less than 20%), non-cardiovascular causes contribute to more than 60% of the burden of hospitalisations^5,29^. Small studies (randomised and observational) have shown that treatment of individual comorbidities such as sleep apnoea^30^, anaemia^31^, AF^32^ and obesity^33^, can reduce hospitalisation and mortality in HF patients. A focus on management of comorbidities is relevant for all types of HF, but particularly for HFpEF^34^ in which no treatment has been uniformly demonstrated to have prognostic benefit. Previous data from our centre has also demonstrated that in 48% of HF patents reviewed in a joint cardio-renal MDT meeting reduced the need for further cardiology or renal outpatient follow-up^35^. However, there is significant variation and disparity in the management of heart failure and associated co-morbidities depending on the geographical location of the patient and health care professional^36^. Specialists frequently focus on a single disease entity, potentially with a deleterious effect on other organ/ systems (one such example is the occurrence of worsening renal function in patients with HFrEF during decompensated HF, prompting cessation of renin-angiotensin aldosterone inhibitors and in turn leading to worsening HF outcomes).

Disease management programs incorporating MDT input from dietician, social worker, physical therapist, and pharmacist have shown a significant reduction in thirty-day readmissions rates^37^, however we note the variable definition of the constitution of a MDT and variable effects of MDT input into HF outcomes^38^. Members of the MDT have included HF specialist nurses, HF consultants, pharmacists, dieticians, social care workers, physiotherapists, palliative care specialists and psychologists. The largest study of MDT intervention (COACH Study) showed neutral outcomes in terms of HF hospitalisations or mortality^39^. It is possible that the lack of specialist input into intensive management of comorbidities may have diluted the effects of MDT input. The strategy of incorporation of other specialists (nephrologist, diabetologist, geriatrician, chest physician, pharmacologist, palliative care) to address comorbidities simultaneously along with HF specialists, is novel and timely, particularly considering the cardiac, renal and metabolic impact of heart failure therapies such as the SGLT2 inhibitors.

Our service model (Figure 2) exemplifies individualised, patient-centric, holistic care using a consensus approach targeted to the management of HF and comorbidities jointly. The need to address and manage co-morbidities frequently prompts referral to other specialty out-patient clinics. Repeated out-patient clinic attendances are inconvenient to otherwise frail and often less mobile patients. We have demonstrated that a virtual multispecialty MDT is associated with time, travel and cost savings and a significant reduction in inter-specialty referrals/ outpatient clinic attendances.

**Figure 2.**
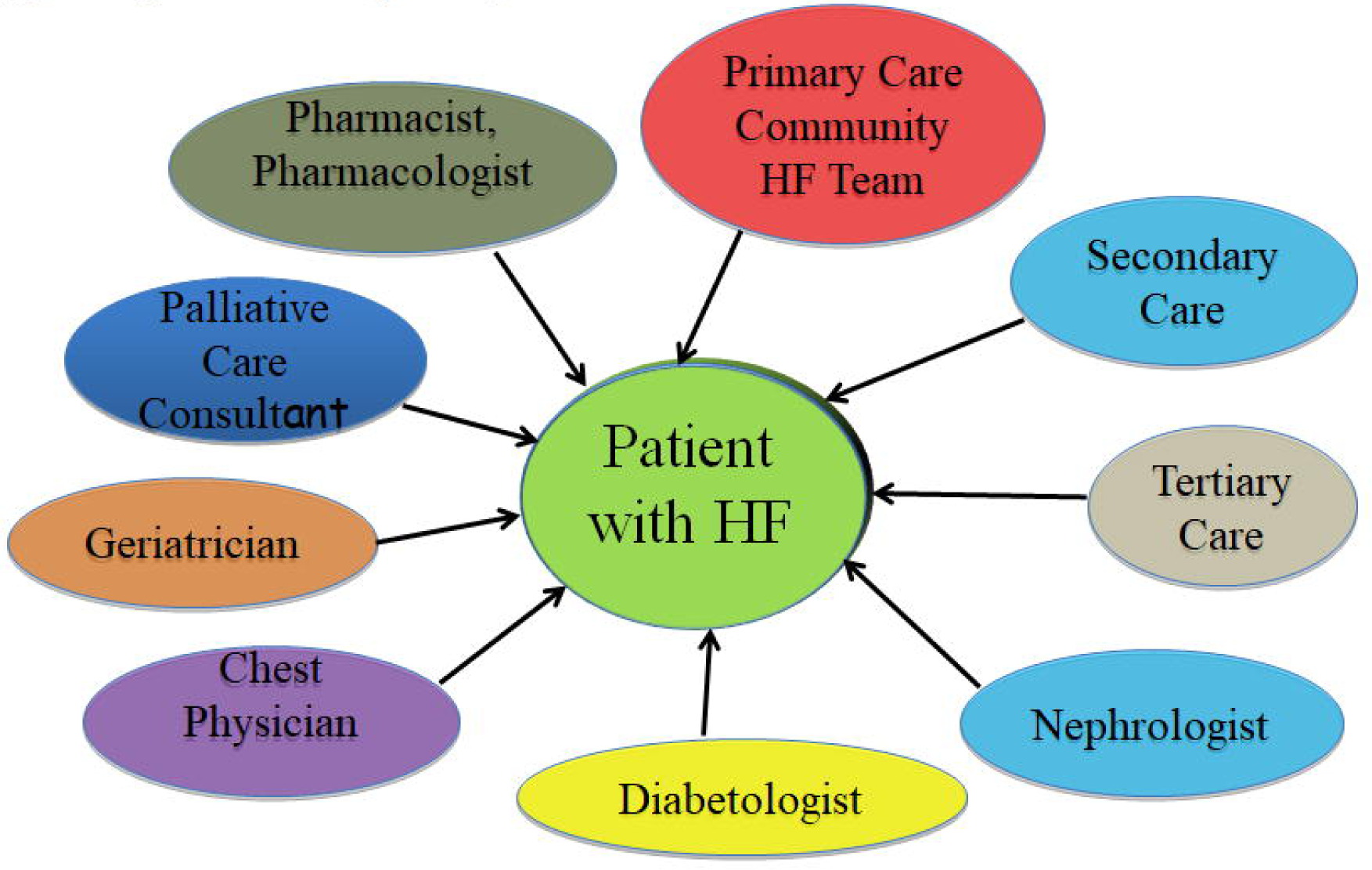
Integrated Multispecialty MDT Model illustrating the team members involved in the MDT meetings

Virtual MDT clinics have been particularly relevant during the COVID-19 pandemic when this particularly vulnerable cohort prefer to avoid travel and minimise the risk of hospital-acquired infection (findings supported by our national HF patient survey regarding the impact of the COVID-19 pandemic upon HF services)^40^. Our virtual multispecialty MDT model importantly ensured that the management of patients with HF continued uninterrupted throughout the COVID-19 pandemic, whilst minimising the need for patients to attend face to face outpatient clinic appointments.

We acknowledge limitations to this study including the observational nature; the efficacy of this service model maybe be best evaluated through a randomised controlled trial. Limitations of the virtual MDT approach also include the potential for miscommunication in view of lack of patient presence during the meeting. This was minimised by spoken and written discussions with the patient prior to and subsequent to the meeting. The model requires robust service planning to ensure attendance of multiple specialists and members of the multidisciplinary team.

In conclusion, we demonstrate that an integrated and virtual multidisciplinary input is associated with improved heart failure outcomes through a reduction in all-cause hospitalisations and clinic attendances and is not only patient-centred but also cost-effective. Application of this model could be considered the gold standard approach in addressing multi-morbidity in patients with heart failure.

## Supporting information

Referral Form

## Data Availability

All data produced in the present study are available upon reasonable request to the authors

## Abbreviations

HF: Heart Failure
MDT: multidisciplinary team
IHD: Ischaemic Heart Disease
AF: atrial fibrillation
HTN: hypertension
CKD: chronic kidney disease
DM: diabetes mellitus
COPD: chronic obstructive pulmonary disease

## Acknowledgements

none

## Data Availability Statement

The data underlying this article will be shared on reasonable request to the corresponding author

