## Supplementary material for "Multispecialty multidisciplinary input into comorbidities in heart failure reduces hospitalisation and clinic attendance": Referral Form

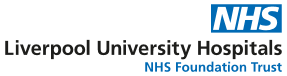
**LIVERPOOL HEART FAILURE MULTISPECIALITY MDT REFERRAL FORM**

| **Patient Details (Print or affix addressograph label)** | | | |
| --- | --- | --- | --- |
| **Surname:** | | **Forename:** | |
| **Address:** | | | |
| **Postcode:** | **DOB:** | | **NHS No:** |

**DIAGNOSIS (please circle) –	HFrEF	HF pEF**

**SPECIALIST INPUT REQUIRED (please circle)**

- **HF Cardiologist –**
- Medication Advice	-	Need for IV Diuretics
- Consideration of Device Therapy	-	Advanced HF / Transplant referral
- Advanced Care Planning
- Other (please specify) ……………………………………………………………………
- **Nephrologist -** Renal diagnosis ……………………………………………………………..
- Cardiorenal syndrome with worsening renal function	- Post AKI advice
- Hyperkalaemia
- Renovascular disease
- Nephrotic or nephritic syndrome
- CKD with anaemia or mineral bone disease or acidosis or resistant hypertension
- Renal transplant or dialysis assessment
- **Diabetologist**
- Poor Diabetes control (Latest HbA1c) …………….. ….Date ……………….
- Advice regarding diabetes medication
- Please specify if	Type 1 DM	Type 2 DM
- Current diabetes medication ……………………………………………………
- **Geriatrician**
- Frequent falls and possible relation to polypharmacy
- Cognitive impairment without diagnosis
- Consideration of ceilings of treatment
- **Chest physician**
- COPD / asthma treatment optimisation, Cor Pulmonale, Interstitial Lung Disease
- Suspicion of sleep apnoea
- Unilateral pleural effusion
- **Palliative Care Consultant**
- Need for out-patient or community palliative services
- Need for hospice care
- **Pharmacist, Clinical Pharmacologist**
- Medication reconciliation, adherance and review of polypharmacy
- Issues regarding availability of medication

Name of Referrer ………………………………………Location …………………Date ……………
